# Predicting Motor Recovery After Stroke: Utility and Limits of Corticospinal Tract Biomarkers

**DOI:** 10.64898/2026.06.16.26355795

**Authors:** Eva S. Rickers, Theresa Paul, Frauke Esser, Lukas Hensel, Elizabeth Rizor, Ellen Binder, Anne K. Rehme, Corinna Ringmaier, Anna Schönberger, Caroline Tscherpel, Christian Grefkes, Scott T. Grafton, Gereon R. Fink, Lukas J. Volz

**Affiliations:** Faculty of Medicine, University of Cologne, and Department of Neurology, University Hospital Cologne, Cologne, Germany; University of California, Santa Barbara, Department of Psychological and Brain Science, Santa Barbara, CA, United States; University of California, Santa Barbara, Institute for Collaborative Biotechnologies, Santa Barbara, CA, United States; University Hospital Cologne, Department of Psychiatry, Köln, Germany; Department of Neurology, University Hospital Frankfurt, Goethe University Frankfurt, Frankfurt am Main, Germany; Institute of Neuroscience and Medicine, Cognitive Neuroscience (INM-3), Research Centre Juelich, Juelich, Germany

**Keywords:** Pyramidal Tracts, Magnetic Resonance Imaging, Machine Learning, Motor Skills, Movement, Rehabilitation, Brain Mapping, Lesion Mapping

## Abstract

**Background:** Corticospinal tract (CST) damage is a major cause of post-stroke motor deficits. However, it remains unclear which estimates of CST damage best predict motor recovery, especially regarding different aspects of motor control. While conventional CST-lesion metrics offer superior feasibility, data-driven machine learning (ML) approaches may better capture patients’ propensity for task-specific recovery with important implication for their use as future clinical biomarkers.

**Methods:** Providing the first direct longitudinal comparison of these approaches based exclusively on CST-lesion patterns, we evaluated six conventional CST-lesion metrics and a voxel-wise ML approach using clinical MRI data from 127 acute ischemic stroke patients. Acute impairment and outcome (>3 months post-stroke) were assessed for basal and complex motor functions. Conventional CST-lesion metrics and ML were used to predict task-specific motor impairment and outcome.

**Results:** All conventional CST-lesion metrics correlated significantly with both acute impairment and motor outcome across motor domains, with metrics weighted for CST narrowing and tract probability performing best. However, predictive performance for unseen patients was low. ML outperformed conventional markers in predicting acute impairment across motor domains and basal motor outcome, but failed to predict complex motor outcome. Topographically, predictive voxels clustered within and above the posterior limb of the internal capsule, with distinct CST subregions associated with basal versus complex motor impairment, consistent with a task-specific somatotopic organization.

**Conclusions:** The predictive utility of CST biomarkers was task- and timepoint-dependent. While ML may improve predictive performance, complex motor outcome remained difficult to predict, likely reflecting greater reliance on distributed cortical reorganization beyond the CST. By revealing task-specific CST subregions, voxel-wise ML provides an anatomically informed foundation for future predictive models. Such future models should combine CST biomarkers with measures of broader motor network integrity to enable individualized prognosis tailored to specific motor domains and recovery stages.

## Background

Corticospinal tract (CST) integrity is among the most promising biomarkers for predicting motor impairment and recovery after stroke.^1–7^ Several metrics have been proposed to quantify structural CST integrity from routine clinical imaging, including different approaches to calculate CST lesion load^1,2,8^ or voxel-wise machine learning (ML).^9,10^ However, the reported amount of explained variance in motor impairment and outcome by distinct conventional CST-lesion metrics drastically varies between 12–70%,^1,2,6,11,12^ hindering clinical implementation. This large amount of variance may arise from several factors such as the choice of CST-lesion metric, the type of motor assessment, and the timing of post-stroke imaging and motor assessment.^2,11,13,14^

While outcome prediction is of high clinical relevance, the few studies that have directly compared different timepoints showed that conventional CST-lesion metrics typically predicted acute impairment far better than outcome.^2,11,15^ Moreover, different aspects of motor function, such as more basal (e.g., grip strength) or more complex motor control (e.g., requiring coordinated multi-joint movement or visuomotor integration), have been hypothesized to rely on distinct recovery mechanisms.^11,13,14,16^ For acute impairment, limited evidence suggests that conventional neuroimaging markers of CST integrity may more strongly relate to measures of complex (e.g., visually guided reaching, Purdue Pegboard Task, Action Research Arm Test (ARAT)) compared to more basal (e.g., grip strength, shoulder abduction finger extension (SAFE) strength) motor function.^11,17^ However, the frequent use of composite^9,10,18^ or a single motor score^2,15^ precludes further task-specific insights. Thus, it remains largely unclear whether conventional CST-lesion metrics feature distinct degrees of predictive potential regarding the recovery and outcome across different aspects of motor control.

Conceptually, such task- and time-dependent differences in predictive performance may arise for several reasons. First, recovery of complex motor function may rely on distributed networks of motor regions, with CST integrity adding little predictive value once CST-independent reorganization occurs.^14,16^ Second, different tract-based metrics may capture distinct anatomical features (e.g., lesion load vs. cross-sectional narrowing vs. tract probability), which might carry varying task-dependent relevance.^6,11,19^ Finally, the precise lesion location within the CST, not merely the overall extent of tract damage, may differentially affect the recovery of more basal and more complex motor functions along two related anatomical axes. Functionally distinct CST sub-tracts arising from primary versus premotor cortical areas may contribute differentially to basal and complex motor control,^19,20^ while the somatotopic organization of the CST also separates more distal (hand and finger) from more proximal (shoulder and arm) representations.^21–23^ These two axes are distinct yet interrelated as basal as well as complex motor control may involve both proximal and distal effectors.^13,14^

While conventional CST-lesion metrics provide clinically feasible proxies of CST damage, they cannot identify specific CST subregions relevant to different functional domains. Here, ML techniques may improve the prediction of motor outcome. Indeed, initial findings from cross-sectional studies suggest that network-level ML approaches may outperform conventional markers of CST integrity in explaining initial motor impairment.^24^ However, similar comparisons for longitudinal predictions of motor outcome from acute baseline imaging are lacking, and ML applied within the CST has not been directly compared with conventional CST-lesion metrics.^9,10^ This raises the question whether conventional or ML-based quantification of CST integrity is superior for predicting post-stroke motor impairment and, more importantly, outcome. Moreover, it remains unclear whether either approach is more favorable for predicting distinct aspects of motor control such as more basal or complex motor function.

To address these questions, we evaluated six conventional CST-lesion metrics and a voxel-wise ML approach applied to CST lesion data extracted from clinical routine MRI in 127 ischemic stroke patients. Acute impairment and outcome at more than 3 months after stroke were evaluated for basal and complex upper-limb motor function.

Based on prior evidence,^24,25^ we hypothesized that ML-based approaches may outperform conventional CST-lesion metrics in predicting motor impairment, particularly with regard to task-specific differences. We assumed that both approaches more accurately predict impairment than outcome,^2,11,25^ given that outcomes can be considered to rely on additional factors such as the reorganization of task-specific motor networks beyond the CST.^12,14,26^ Finally, we anticipated task-related differences to be more pronounced in outcome prediction since the recovery of different functional domains can be expected to rely on task-specific mechanisms of compensation and reorganization beyond the degree of CST integrity.^13,14,19,20^

## Methods

### Data Availability Statement

Data are available from the corresponding author upon reasonable request. This article follows the Strengthening the Reporting of Observational Studies in Epidemiology (STROBE) reporting guideline.

### Participants

131 first-ever ischemic stroke patients were recruited from the Stroke Unit of the Department of Neurology at the University Hospital Cologne, Germany. Inclusion criteria were (1) written informed consent, (2) age between 40 and 90 years, and (3) hemiparesis of the upper limb at hospital admission. Exclusion criteria were (1) bihemispheric stroke, (2) primary intracerebral hemorrhage, (3) contraindications to MRI, and (4) National Institutes of Health Stroke Scale (NIHSS) score >20. The study was approved by the local ethics committee at the University of Cologne and conducted in accordance with the Declaration of Helsinki. A subset of our current patient cohort has recently been published in a previous study assessing region-to-region disconnections and motor recovery.^14^

### Imaging and behavioral assessment

MRI scans were obtained as part of the standard clinical protocol using either a 1.5 T Philips (Philips, Guildford, United Kingdom) or a 3.0 T Siemens scanner (Siemens Medical Solutions, Erlangen, Germany). For each patient, both basal and complex upper extremity (UE) motor function were assessed at hospital admission (impairment) and at a follow-up examination more than 3 months after stroke (outcome). More basal motor function was assessed via relative grip strength, calculated as the ratio of mean grip force across three vigorimeter trials of the affected to the unaffected hand. More complex aspects of motor control were quantified using the Action Research Arm Test (ARAT), which assesses activities of daily living involving multi-joint movements and visuomotor integration.^27^ Three patients were excluded due to poor MRI quality, and one patient was excluded due to incomplete motor assessments. The final sample comprised 127 patients, of whom 92 patients (72.4%) completed the follow-up examination.

### Image preprocessing

Image preprocessing and further analyses were conducted using Python (version 3.11.5), including NumPy, pandas, SciPy, scikit-learn (version 1.5.2), matplotlib, seaborn, nibabel, statsmodels, and FSL (version 6.0.7.6). Lesion masks were manually drawn on clinical diffusion-weighted images (DWI), or fluid-attenuated inversion recovery (FLAIR) images, or T2-weighted images using MRIcron,^28^ depending on image quality, and were verified by a certified neurologist. Lesions were binarized and, if located in the right hemisphere, flipped along the midsagittal axis, resulting in left hemispheric lesions across all patients.

Image normalization was performed by co-registering skull-stripped DWI images and lesion masks to individual T1- or T2-weighted images and subsequently normalizing them to MNI space. The MNI-icbm152 6^th^ generation symmetric template was used as the standard template (https://nist.mni.mcgill.ca/mni-icbm152-non-linear-6th-generation-symmetric-average-brain-stereotaxic-registration-model/).^29^ All spatially normalized lesion masks were visually inspected to ensure accurate spatial transformation to MNI space.

### CST template

The CST atlas by Chenot and colleagues^30^ (https://www.gin.cnrs.fr/en/tools/pyt-atlas/) was used to define the CST. A binary CST template was generated by thresholding the probabilistic CST map at 20%, retaining voxels containing CST fibers in ≥20% of individuals, in line with previous studies.^31^

To account for varying probabilities of voxel-wise CST inclusion across individuals, the probabilistic CST template was thresholded at ten distinct levels between 10% and 99% (in 10% steps, last step 9%) in line with previous work.^1^ The resulting ten binary CST templates were summed to create a map with ten discrete intensity levels (I), where I = 1 corresponds to 10% and I = 10 to 99% probability of CST inclusion.

### Conventional CST-lesion metrics

Lesion volume (regardless of CST localization; Figure 1A) and the following six distinct conventional CST-lesion metrics were calculated: (B) CST**-**lesion volume, (C) raw maximum cross-sectional CST-lesion area, (D) maximum cross-sectional CST-lesion area weighted by the cross-sectional CST-area of the corresponding slice,^32^ (E) Feng-overlap,^2^ (F) raw Zhu-overlap,^1^ and (G) weighted Zhu-overlap^1^ (Figure 1).

**Figure 1:**
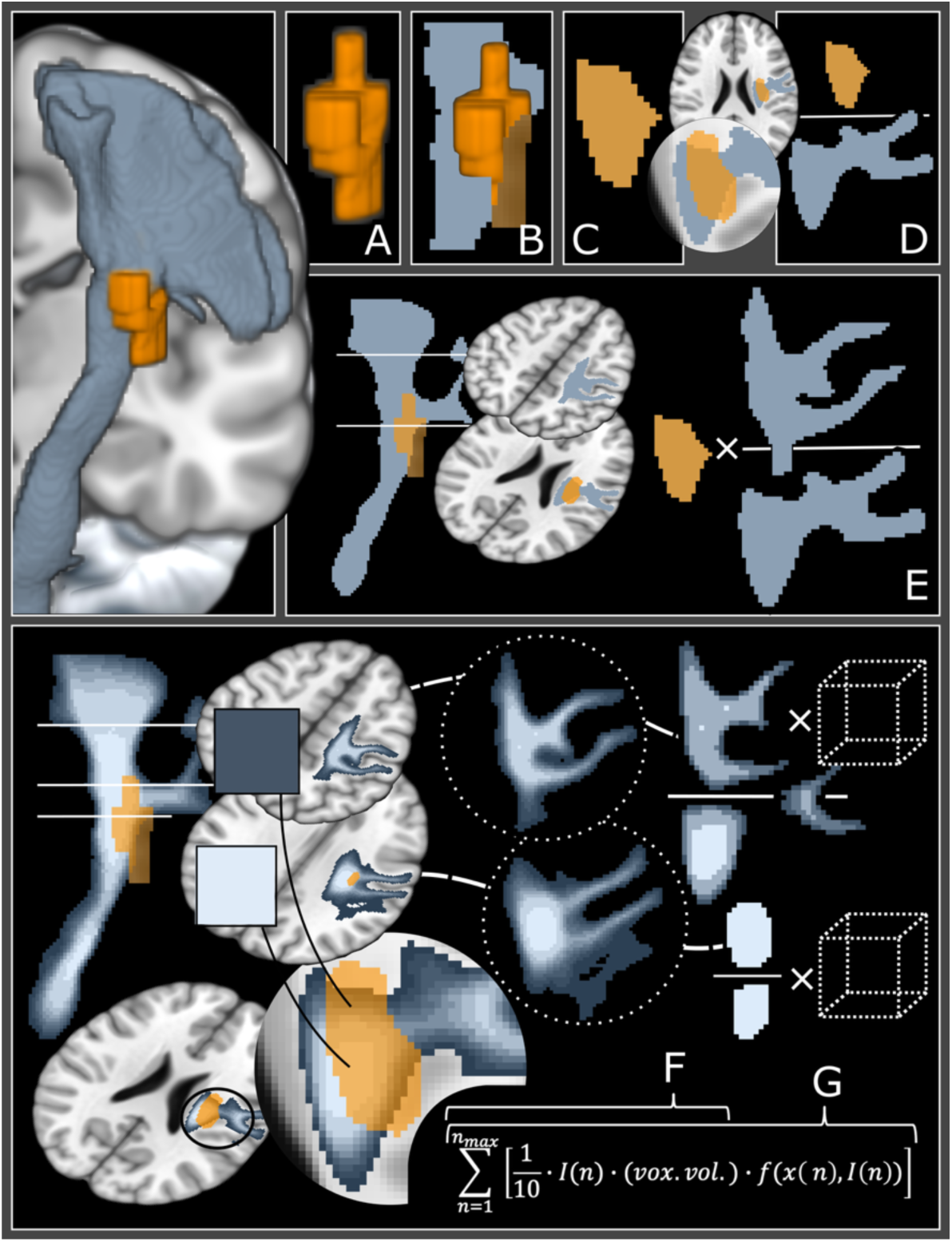
Conventional corticospinal tract lesion metrics. (A) lesion volume, (B) CST-lesion volume; (C) raw maximum CST-lesion area; (D) weighted maximum lesion-CST-area, which is weighted per cross-sectional CST-area of corresponding slice; (E) Feng-overlap;^2^ (F) raw Zhu-overlap^1^; (G) weighted Zhu-overlap^1^

Feng-overlap was calculated as a weighted CST-lesion metric in accordance with Feng et al.^2^ For each axial slice, the lesion overlap with the CST was weighted by the ratio of the maximum CST cross-sectional area to the CST cross-sectional area at that slice.

Raw and weighted Zhu-overlap were computed based on the approach proposed by Zhu et al.^1^ Accordingly, the raw Zhu**-**overlap was defined as the sum of lesioned CST-voxels, each multiplied by its intensity level (ranging from 1–10), representing their probability (in 10% steps) of being located within the CST.^1^ For the weighted Zhu-overlap each voxel was additionally multiplied by a weighting factor accounting for the narrowing of the descending CST. This weighting factor was calculated individually for each slice and intensity level *(I)* as the ratio of the number of voxels ≥ *I* on the slice containing the highest such count to the number of voxels ≥ *I* on the respective voxel’s slice.^1^

Spearman correlation analyses were conducted for lesion volume and each of the six CST-lesion metrics with measures of impairment and outcome for both basal and complex motor function, and were corrected for multiple comparisons.^33^

### Leave-one-out (LOO) regression analyses

To facilitate comparison with the support vector regression (SVR) model, iterative leave-one-out (LOO) regression analyses were conducted for each conventional CST**-**lesion metric.^24^ For each iteration, one patient was excluded, and the regression model was trained on the remaining set of patients to predict the left-out case, thereby providing a cross-validated estimate of predictive performance on unseen patients. Z**-**score standardization of target variables was applied to mirror the preprocessing of the SVR models.^34^ For 127 patients with impairment data, the LOO approach yielded 127 cross-validated predictions, each based on a distinct regression model. The predictive performance of each conventional CST-lesion metric was then quantified as the Spearman correlation and the coefficient of determination (*R²*) between the 127 cross**-**validated predictions and the observed motor scores.

### Support vector regression analysis

For voxel-wise ML-based prediction of basal and complex motor function, support vector regression (SVR) was implemented using a nested cross-validation approach with Scikit-learn (version 1.5.2; see Figure S1 for a schematic overview).^34^ Model performance was evaluated in the outer loop using leave-one-out cross-validation (LOOCV) and quantified as the root mean squared error (RMSE) across all held-out predictions. In addition, the coefficient of determination (R²) and the Spearman rank correlation between predicted and observed scores were computed to assess SVR performance relative to conventional CST**-**lesion metrics. Hyperparameter tuning was conducted in the inner loop using an N/5**-**fold grid**-**search cross**-**validation with either a linear or a nonlinear (radial basis function or polynomial) kernel, in line with previous work.^14,35^

A total of 51,568 lesioned CST voxels were used as binary input features (0 = non-lesioned, 1 = lesioned) and sorted by lesion frequency across patients. Since voxels lesioned in only a few patients carry limited variance and may yield unstable feature weights, and a high feature-to-sample ratio necessitates dimensionality reduction, SVR models were trained across varying subsets of the most frequently lesioned CST voxels, ranging from 0.01% (N = 5) to 15% (N = 7735) of all features.^36,37^ The optimal model across the feature space was identified via the lowest RMSE (Figure S2).

Z-score standardization of target variables was applied.^34^ To estimate feature importance in nonlinear SVR models, we averaged the Lagrange multipliers (αᵢ) from the support vectors in the decision function across folds, serving as an approximation of voxel-wise feature weights.^38^ Averaged feature weights from linear and nonlinear SVR models were significantly correlated (all r≥0.77, p<0.001), indicating consistent feature representations across model types and supporting the interpretability of feature patterns derived from nonlinear models (see Table S3).^39^

### Localizing predictive CST-voxels

To visualize the contribution of individual CST voxels to the prediction of motor impairment and outcome, voxel-wise feature weights were mapped to MNI standard space. To enable comparability across models and tasks, voxel-wise feature weights were z-standardized relative to a permutation-based reference distribution in accordance with previous work.^14,35^ This reference distribution reflects the range of feature weights assuming the lack of a systematic relationship between CST lesion patterns and motor performance. 1,000 permutations, including SVR model retraining, were performed. For each permutation, target variables were randomly reassigned to input features. Color-coded maps of voxel-wise feature weights were then used to visualize the spatial distribution of voxels contributing to model predictions (Figure 5). Feature weights were visualized for voxels in which lesion presence indicated poorer motor performance.^14,37^

To compare voxel-wise predictive relevance regarding more basal and complex motor control, we computed contrasts between SVR-derived feature weights using a permutation-based approach (see Supplemental Methods for a detailed description).^40,41^ For each permutation, behavioral labels were randomly reassigned and SVR models were fitted separately for basal and complex motor function, yielding voxel-wise permutation weight maps. These maps were subtracted to obtain permutation-based difference maps. Across permutations, the mean and standard deviation of the voxel-wise difference distribution were computed and used to standardize the observed difference between the true models for basal and complex motor function. This procedure yielded a voxel-wise contrast metric that reflects the scale-invariant relative difference in predictive relevance between more basal and complex motor function. The resulting standardized difference map was visualized in MNI space to illustrate spatial patterns of task-specific contributions.

## Results

A total of 127 patients (mean age 66.07 ±12.24 years, days since stroke: median 1, IQR = 3) were included in the analyses of motor impairment. 92 patients also completed outcome assessment in a follow-up session (months since stroke: median 4.20, IQR = 1.29). Patients exhibited a wide range of initial motor impairment and highly heterogeneous recovery trajectories across both basal and complex motor domains (see Figure 2 for individual behavioral data). Across the cohort, lesions covered the entire territory of the middle cerebral artery, with the greatest overlap in subcortical areas traversed by the corticospinal tract. Within this pathway, the internal capsule was the most frequently affected region, and some lesions extended further downward into the brainstem (Figure 2).

**Figure 2:**
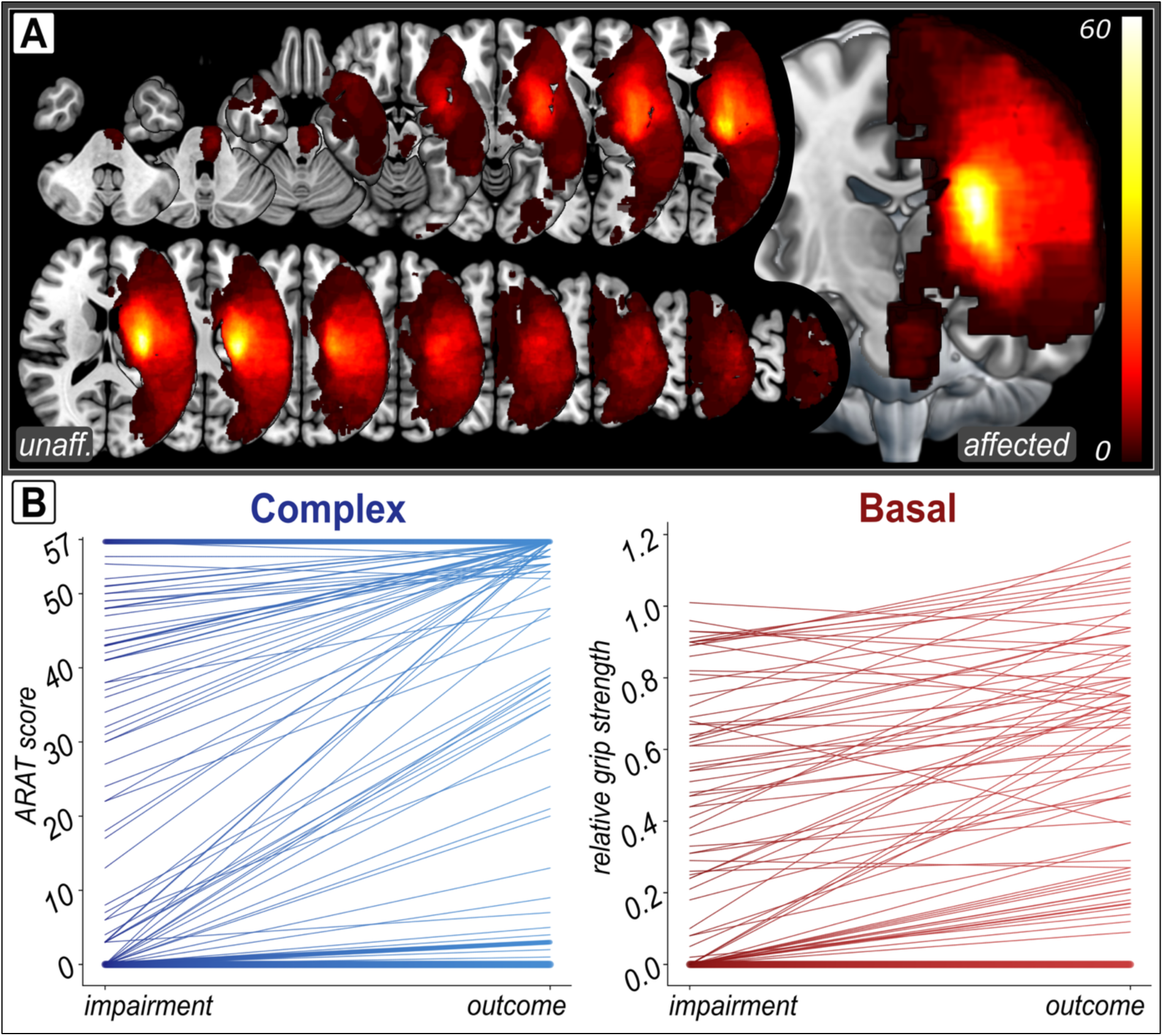
Lesion distribution and motor recovery trajectories. (A) Overlay of all normalized lesion masks in standard space, with right-hemispheric lesions flipped to the left hemisphere. Lesion coverage spanned the entire territory of the middle cerebral artery, concentrating in subcortical regions traversed by the corticospinal tract, and extending into the brainstem in a subset of patients. Color intensity reflects the number of patients with a lesion at each voxel. The most frequently lesioned voxel was affected in 60 patients. (B) Individual recovery trajectories from acute impairment to outcome, shown separately for more complex (ARAT, left) and more basal (relative grip strength, right) motor function. We observed substantial heterogeneity in both initial impairment and recovery for both motor domains.

### Conventional CST-lesion metrics

Lesion volume and all conventional CST-lesion metrics significantly correlated with impairment of both basal and complex motor function (Figure 3A, all p < 0.05, FDR**-**corrected for multiple comparisons). Regarding motor outcome, significant correlations were observed for all CST-lesion metrics but not for lesion volume.

**Figure 3:**
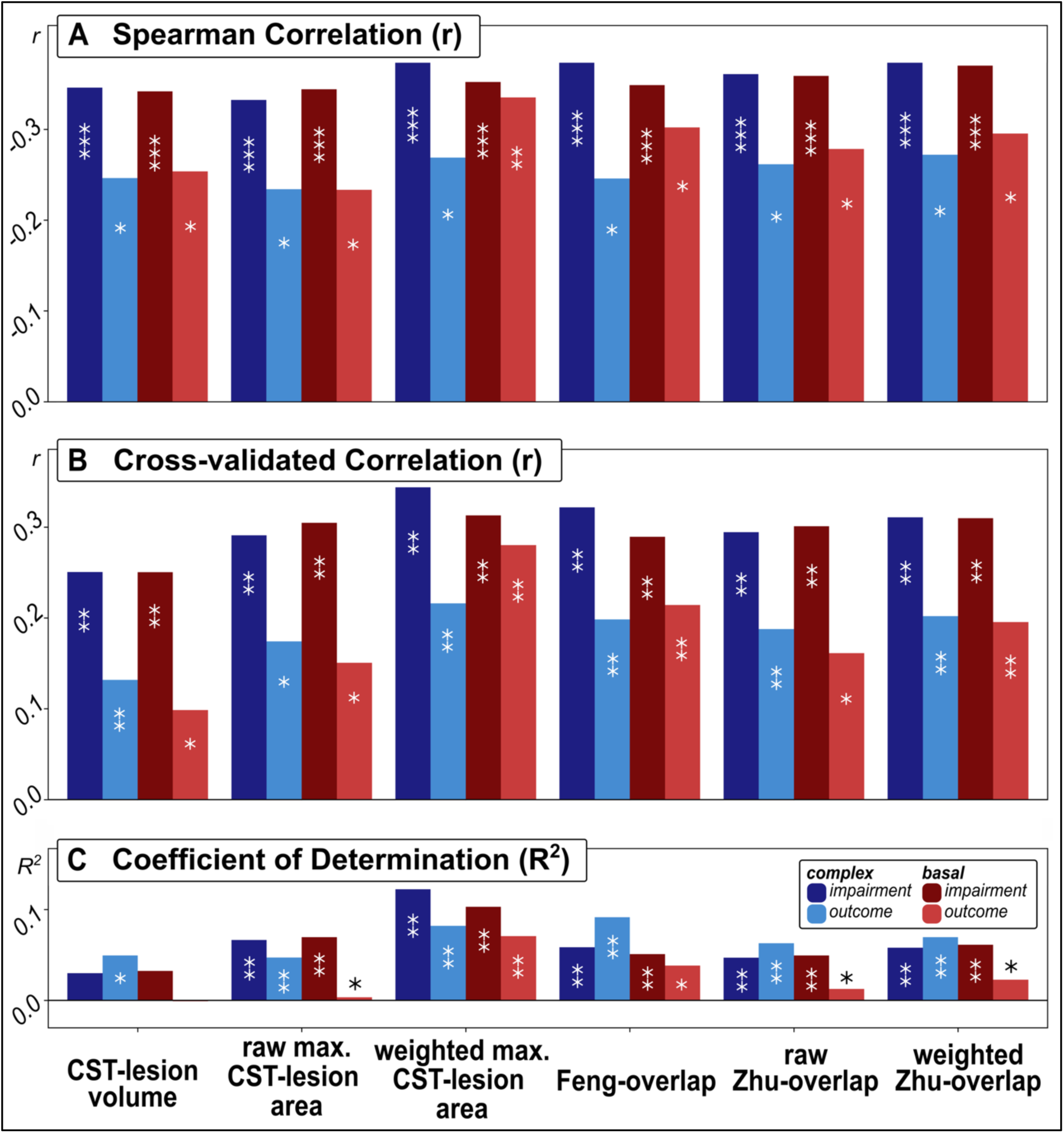
Correlations and predictive performance of conventional CST-lesion metrics. (A) Spearman correlation coefficients between CST-lesion metrics and motor scores, (B) leave-one-out cross-validated Spearman correlation coefficients and (C) leave-one-out cross-validated R² for CST-lesion metrics and impairment (darker shades) or outcome (lighter shades) for more basal (red; relative grip strength) and complex (blue; ARAT) motor function. Asterisks indicate FDR-corrected significance (* p < 0.05, ** p < 0.01, *** p < 0.001). Correlations and LOOCV-predictions were consistently stronger for impairment than outcome. Of note, weighted CST-lesion metrics outperformed unweighted variants, with weighted maximum CST lesion area showing the strongest associations. LOOCV performance was substantially lower across all CST-lesion metrics, indicating limited generalization to unseen patients. For exact correlation coefficients, performance metrics, and p-values, see Table S1.

Overall, conventional CST-lesion metrics demonstrated stronger associations with motor impairment than outcome (Figure 3, Table S1). Notably, CST-lesion metrics accounting for the narrowing of the descending CST via a weighting factor (see Figure 1D, 1E, 1G) or for the probability of a lesioned voxel belonging to the CST (see Figure 1F) yielded higher correlations with behavioral scores compared to unweighted (raw) overlap measures.

To enable direct comparison of predictive utility between conventional CST**-**lesion metrics and ML-based approaches, we computed cross-validated predictions using conventional CST-lesion metrics. Cross-validated performance for unseen patients was low across all conventional CST-lesion metrics (Figure 3B, C). Differences between CST**-**lesion metrics mirrored the results of the correlation analysis: performance was (i) higher for impairment than outcome and (ii) higher for weighted than unweighted CST-lesion metrics, with weighted maximum CST lesion area performing best overall.

### Support vector regression (SVR)

The best-performing SVR model for predicting motor impairment included 10% of the most frequently lesioned CST voxels for both basal and complex motor function. Conversely, outcome prediction of more basal motor function only required 2% of the most frequently lesioned CST voxels (Figure S2). No SVR model significantly predicted complex motor outcome. When predicting unseen patients was successful, SVR explained a higher proportion of variance than the best conventional CST-lesion metrics (performance metrics summarized in Figure 4 and Table S2).

**Figure 4:**
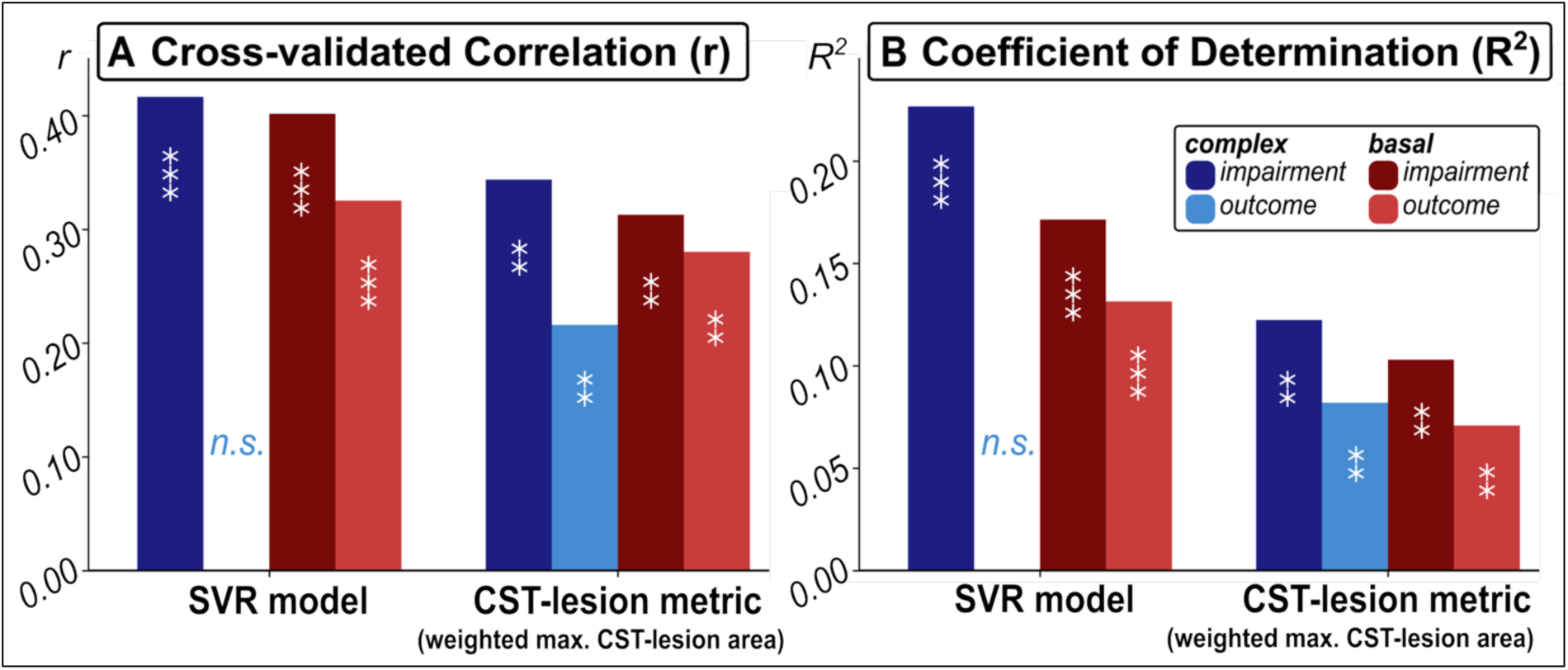
Performance metrics of the support vector regression (SVR) analysis and best conventional CST-lesion metric. The best SVR model for predicting motor impairment used 10% of the most frequently affected voxels for both more basal and complex motor function. In contrast, the best model for predicting more basal motor outcome only included 2% of features (asterisks indicate significance at ** p < 0.01 and *** p < 0.001). Of note, none of the tested SVR models significantly predicted complex motor outcome. (A) Cross-validated correlation coefficients (Spearman r) and (B) Coefficients of determination (R^2^) were higher for significant SVR models compared to the best CST-lesion metric, indicating superior predictive performance.

The most predictive voxels for both basal and complex motor impairment clustered in highly overlapping regions within and just above the posterior limb of the internal capsule (PLIC), consistent with areas of dense fiber convergence and frequent lesion occurrence (Figure 5). For basal motor outcome, predictive voxels were similarly concentrated at this CST bottleneck, within and just above the PLIC, outlining this region as a key determinant of both acute impairment and longer-term basal recovery. Despite the spatial overlap in predictive regions, directly comparing voxel-wise contributions revealed distinct CST subregions differentially informative with regard to more basal versus complex motor impairment (Figure 6). Voxels predictive of more basal impairment clustered in dorsolateral CST portions. Conversely, voxels primarily informing the prediction of complex motor function were located in ventromedial segments. This dorsolateral–ventromedial gradient was consistent across consecutive axial slices, suggesting a spatial subdivision. Of note, this spatial topography aligns with the known somatotopic gradient of the CST,^21–23^ where dorsolateral segments correspond to distal hand and finger musculature underlying grip strength, and ventromedial segments relate more strongly to proximal arm control which, in contrast to distal grip strength, is additionally engaged during ARAT performance.

**Figure 5:**
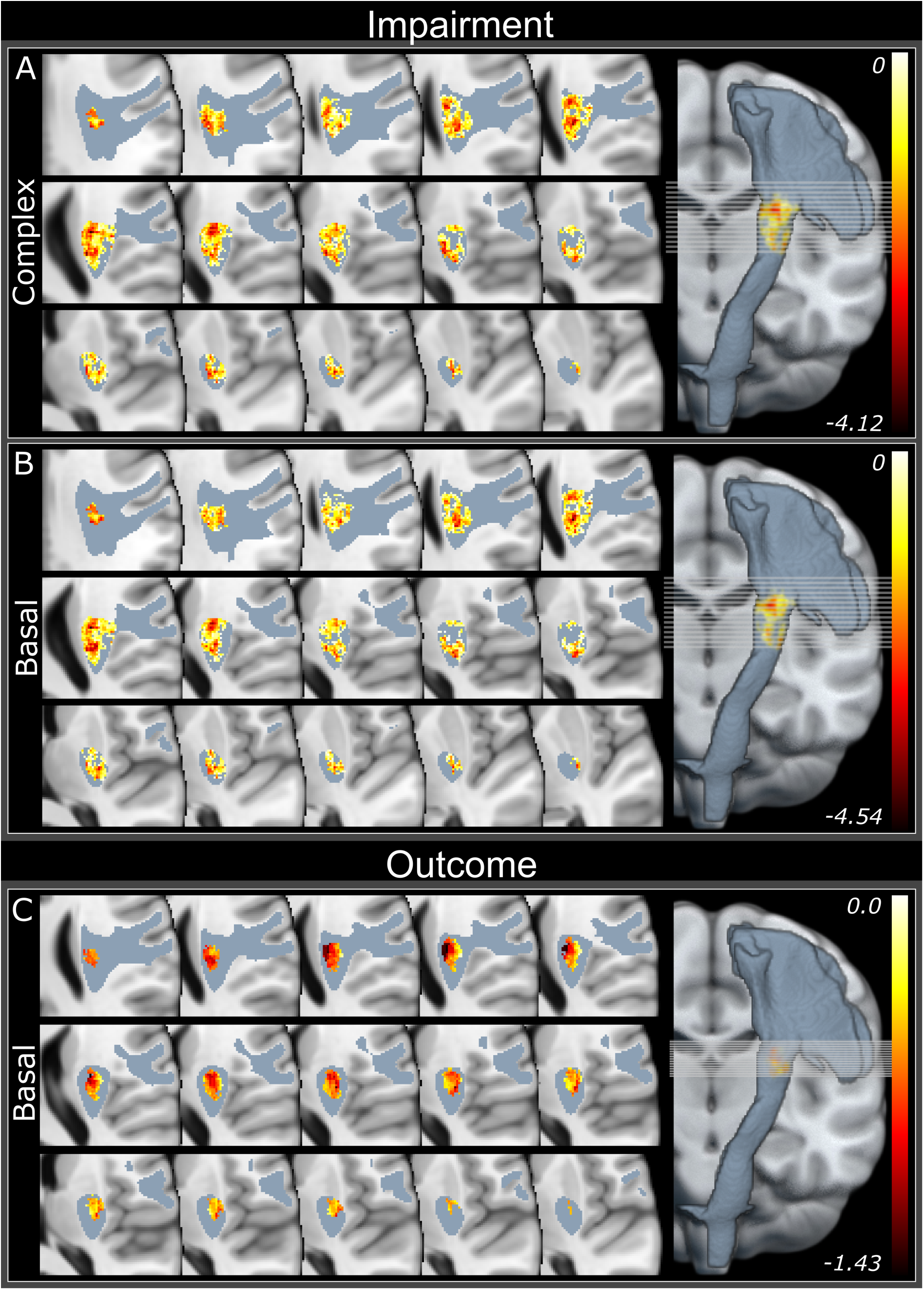
Voxel-wise contributions to the prediction of motor function. Feature weights for SVR models indicative of (A) complex motor impairment, (B) more basal motor impairment, and (C) more basal motor outcome. Darker colors indicate stronger associations between voxel damage and impaired motor function (see color bars). Predictive voxels for both motor domains and timepoints clustered predominantly within and just above the posterior limb of the internal capsule and adjacent superior CST segments, highlighting this fiber bottleneck as a key determinant of acute impairment and basal outcome.

**Figure 6.**
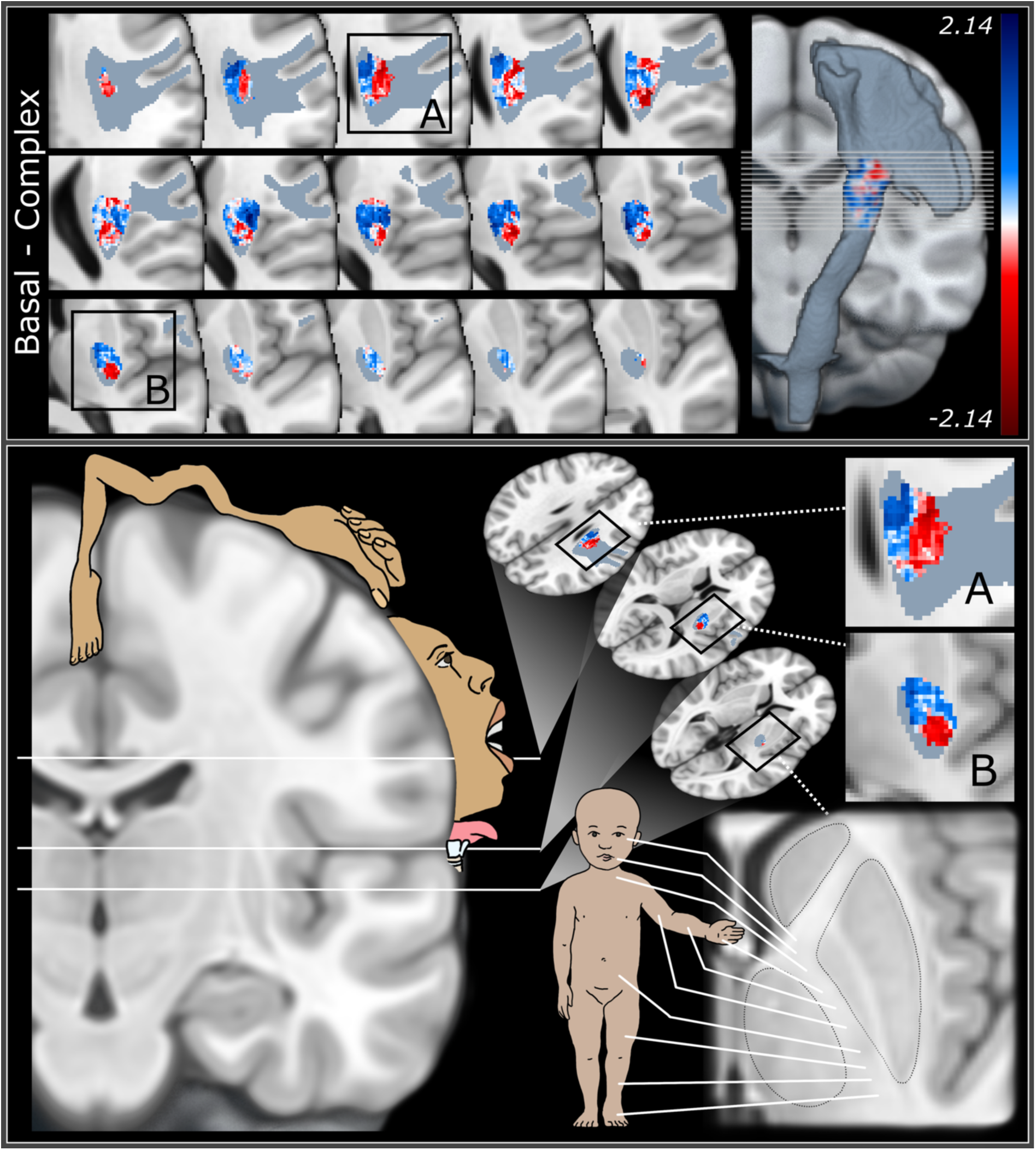
: Dorsolateral–ventromedial gradient of CST voxels indicative of basal versus complex motor impairment. Top panel: voxels primarily indicative of more basal motor impairment (grip strength; red) clustered in dorsolateral CST regions, while voxels primarily contributing to the prediction of complex motor impairment (ARAT; blue) were located in more ventromedial segments. Bottom panels: anatomical context provided by the CST’s somatotopic gradient at the motor cortex (left) and PLIC (right), where medial cortical areas and anterior CST segments govern proximal arm control and lateral cortical areas and posterior CST segments govern distal hand function. This gradient directly maps onto the motor demands of our tasks: ARAT performance additionally requires proximal arm movements (complex, blue), whereas grip strength relies on distal muscle control (basal, red).

### Demographic variables

While age and time elapsed since stroke have previously been reported to explain variance in motor recovery,^42^ neither showed significant correlations with motor function in our patient cohort (*age and impairment:* basal: *Spearman r*=0.07, p=0.50; complex: *Spearman r*=0.08, p=0.35; *age and outcome:* basal: *Spearman r*=0.06, p=0.56; complex: *Spearman r* =0.10, p=0.37; *time since stroke and outcome:* basal: *Spearman r*=-0.06, p=0.56; complex: *Spearman r* =0.09, p=0.42). Of note, these results align with previous findings on CST-lesion metrics in which demographic variables did not significantly contribute to multivariable models. ^6,32^ For example, Liew et al. found that brain-predicted age difference, a surrogate marker of brain aging, accounted for only 4 % of the effect of lesion-CST-overlap on sensorimotor outcomes.^43^

## Discussion

### Prediction of motor impairment after stroke

In line with the well-established importance of CST integrity for motor control after stroke, all CST-integrity metrics were significantly associated with motor impairment (Figure 3, Figure 4). In particular, weighted CST-lesion metrics (Figure 1D–G) demonstrated stronger correlations with motor impairment than non-weighted metrics, supporting the utility of metrics reflecting the number of transected CST fibers, in line with prior studies.^1,2,6,8^ Notably, our results suggest that the weighted maximum CST-lesion area may represent a viable biomarker candidate given its relative simplicity and competitive predictive performance (Figure 1D). Somewhat surprisingly, more complex alternatives, such as the Feng- and Zhu-overlap, did not further improve predictions in our patient cohort (Figure 3). Crucially, when evaluating cross-validated predictive performance on unseen patients, the utility of all conventional CST-lesion metrics was limited, with weighted metrics still performing best. While previous studies have established the value of conventional CST-lesion metrics, they largely relied on standard correlation or regression analyses.^6,11,15^ Thus, our current findings highlight that such approaches capture group-level associations but may hold limited predictive utility for individual, unseen patients, which is paramount for future clinical application. This raises the question of whether higher-dimensional ML approaches may improve the prediction of task-specific motor impairment and patient-level outcomes. Indeed, our ML approach substantially improved the prediction of both basal and complex motor impairment compared to conventional CST-lesion metrics (Figure 4), consistent with prior work demonstrating that data-driven models may better capture acute stroke deficits.^4,24,37^ However, from a clinical perspective, the true utility of an imaging biomarker lies in its ability to predict future outcome and thereby guide personalized therapeutic applications.

### The challenge of outcome prediction

Our results confirm the relevance of CST integrity for motor outcome, as all conventional CST-lesion metrics were significantly correlated with motor outcome (Figure 3). Conceptually, this is consistent with the notion that residual CST integrity is not only critical for the degree of initial motor impairment but that the CST may also serve as structural motor reserve, enabling subsequent functional reorganization and recovery.^26,44^ Accordingly, functional reorganization has been shown to partially depend on the extent of CST-damage.^26^

Then again, all conventional CST-lesion metrics were more closely related to acute motor impairment than outcome and their cross-validated performance in predicting outcome for unseen patients was poor. This attenuation may reflect effects of motor network reorganization that increasingly shape motor control beyond the degree of CST damage.^12,13,16,45^ Despite the fact that residual CST fibers may serve as structural substrates through which reorganized motor networks may enable motor recovery,^12,26^ CST-lesion metrics cannot capture the degree to which remaining CST fibers are functionally recruited. Moreover, compensation may additionally involve alternative descending motor routes, such as extrapyramidal tracts or contralesional non-crossing CST fibers.^13,19^ Together, these processes may attenuate CST–behavior relationships over the course of recovery and thereby limit the predictive value of CST-lesion metrics for motor outcome.

When evaluating whether higher-dimensional modeling could overcome this limitation, our longitudinal ML approach showed that outcome prediction based on CST lesion information was feasible for basal but not complex motor control (Figure 4). Hence, our findings suggest that the added value of high-dimensional voxel-wise modeling may be strongly time- and task-dependent.

### Anatomical features of task-specific motor impairment

Prediction of motor impairment relied on voxels located in a region of densely packed fibers within and just above the posterior limb of the internal capsule (PLIC; Figure 5). This predictive signal was spatially focal rather than distributed along the entire CST. These findings highlight the critical role of this well-known CST bottleneck. This is corroborated by prior work highlighting the prognostic value of CST integrity measured at the PLIC using fractional anisotropy (FA)^46,47^ and support vector machine (SVM)-based classification models.^9^

Beyond this shared reliance on CST bottleneck regions, our ML approach revealed a task-specific spatial gradient of predictive voxels within the CST (Figure 6). Given that our measure of complex motor function included shoulder and proximal arm movements, whereas our measure of basal motor control relied more on distal hand function, the spatial transition revealed by our ML approach appears anatomically plausible. In particular, the distribution of voxels followed the somatotopic organization of the CST (Figure 6): At the cortical level, medial regions have been shown to correspond to more proximal (e.g., shoulder) and lateral regions to more distal arm control (e.g., hand and wrist).^22,23^ As these fibers descend into the internal capsule, this organization is preserved but topographically rotated such that proximal fibers occupy more ventromedial segments, while distal fibers are located more dorsolaterally.^21^ This proximal-to-distal gradient captured by our ML approach directly mirrors the early excitation mapping of the internal capsule in non-human primates.^21^

An additional explanation for this task-specific gradient may lie in the contribution of different CST sub-tracts. Pre- and supplementary motor fibers (e.g., originating from PMd, PMv, SMA) are thought to be located more anteriorly within the PLIC than primary motor cortex (M1) projections and may contribute more strongly to complex motor functions such as inter-joint coordination and visuomotor integration.^3,31,48,49^ We recently showed that while M1 sub-tracts were associated with both basal and complex motor tasks, premotor sub-tracts were more closely linked to complex motor function.^20^ In contrast, basal motor control may more strongly rely on posterior fibers. For example, Schulz and colleagues found that fractional anisotropy in the PLIC for M1 and PMd sub-tracts, but not for anterior tracts like PMv or SMA, significantly correlated with grip strength.^3^ This nicely aligns with early case studies showing increased activation of M1 and PMd during attempted hand grip following CST disruption.^48^

Taken together, these findings suggest that ML-based modeling improves predictive performance particularly for acute impairment and captures spatial variations in the relevance of CST damage that is not reflected by conventional low-dimensional lesion markers.

### Can machine learning improve predictions in clinical settings?

Previous ML studies have suggested that data-driven models may better capture stroke severity.^4,24,37^ However, they typically featured cross-sectional designs at the acute^4,37^ or chronic^24^ stage, often incorporating whole-brain features or broad regions of interest. To the best of our knowledge, this is the first study to provide a direct, longitudinal comparison between conventional CST-lesion metrics and voxel-wise ML models based solely on CST-lesion patterns. This approach allowed us to compare the predictive value of fine-grained spatial topography against low-dimensional metrics for distinct aspects of motor control. Through this direct comparison, our results revealed that the added value of voxel-wise ML is strongly time- and task-dependent as it failed to predict complex motor outcomes.

An explanation for this dissociation may lie in task-specific reorganization. For example, grip strength, which relies on distal muscles, may depend almost exclusively on specific CST fibers,^3,20^ hindering the compensation via alternative routes and tying its recovery more closely to CST damage. In contrast, recovery of more complex motor function might additionally depend on distributed cortico-cortical networks enabling visuomotor integration and inter-joint coordination.^12,14,17,50^ Consequently, CST-lesion load may not capture all relevant mechanisms facilitating recovery of complex motor control.^3,12,50^

Another potential limitation of ML models may arise from their susceptibility to overfitting to acute lesion features.^25,37^ By exploiting fine-grained spatial variations, ML models may be biased to more closely reflect acute impairment rather than outcome.^10,25,37^ Because low-dimensional conventional CST-lesion metrics provide a single global estimate of tract damage, they are inherently less prone to the severe overfitting seen in high-dimensional models.^4,25,37^

From a clinical perspective, these findings suggest that ML-based modeling of CST damage may offer particular advantages when predicting motor functions closely linked to focal corticospinal injury, whereas prediction of motor recovery remains challenging. Future predictive models may therefore benefit from task- and time-specific adaptation that integrates CST-based structural markers with measures of broader motor network integrity.

### Limitations

Given that we used a nonlinear SVR, one may argue that feature weights should not be interpreted as proxies for the voxel-wise contributions to the prediction of behavioral endpoints. Then again, when repeating the analyses using a linear SVR, the resulting features were highly correlated with the nonlinear features (see Table S3 for further details), corroborating the interpretability of voxel-wise contributions to behavioral predictions.^36^

Moreover, voxel-wise feature weights are influenced by lesion frequency. Ideally, for maximum discriminative power, each voxel would be lesioned in roughly 50 % of patients to balance variance between lesioned and non-lesioned cases. While this would theoretically optimize voxel-level inference, it would neglect the real-world distribution of stroke lesions arising from the vascular anatomy of the human brain.^36^

Finally, the modest size of our follow-up sample (n = 92) may limit statistical power for outcome prediction, particularly regarding the detection of more distributed effects expected for complex motor function. However, the successful prediction of basal motor outcome and the fact that comparable sample sizes have yielded robust predictions in related previous work^14^ suggest that sample size alone is unlikely to account for the lack of significant ML prediction for complex motor function.

### Conclusions

We here provide the first direct comparison of conventional and ML-based markers of CST-lesion load for the longitudinal prediction of distinct aspects of motor function. Conventional CST-lesion metrics were indicative of both motor impairment and recovery for more basal and complex aspects of motor control, highlighting the robustness of low-dimensional anatomical summaries to heterogeneous task-dependent recovery processes. In contrast, voxel-wise SVR models enhanced predictive performance in a time- and task-dependent manner, revealing anatomically distinct CST subregions that contribute to more basal versus more complex motor impairment. Of note, ML-based longitudinal prediction was only feasible for the outcome of basal but not complex motor control. This pattern likely reflects increasing contributions of distributed cortico-cortical motor networks and variability in the functional recruitment of residual CST pathways during recovery, which cannot be captured by metrics of structural CST integrity alone.

Taken together, our results outline the task- and time-dependent limitations of using CST damage as a clinical biomarker for motor recovery. Moreover, our current findings provide a foundation for future predictive models integrating CST-based structural markers with measures of distributed motor network integrity to further refine individualized prognosis and inform personalized rehabilitation strategies after stroke.

## Data Availability

Data are available from the corresponding author upon reasonable request.

## Acknowledgments

None.

## List of Supplemental Material

Supplemental Methods

Figures S1, S2

Tables S1–S3

## Nonstandard abbreviations and acronyms

ARAT: Action Research Arm Test
CST: Corticospinal Tract
DWI: Diffusion-Weighted Imaging
FA: Fractional Anisotropy
FLAIR: Fluid-Attenuated Inversion Recovery
IQR: Interquartile Range
LOO: Leave-One-Out
LOOCV: Leave-One-Out Cross-Validation
ML: Machine Learning
M1: Primary Motor Cortex
MRI: Magnetic Resonance Imaging
NIHSS: National Institutes of Health Stroke Scale
PLIC: Posterior Limb of the Internal Capsule
PMd: Dorsal Premotor Cortex
PMv: Ventral Premotor Cortex
RMSE: Root Mean Squared Error
SMA: Supplementary Motor Area
SVR: Support Vector Regression

## References

1. Zhu LL, Lindenberg R, Alexander MP, Schlaug G. Lesion load of the corticospinal tract predicts motor impairment in chronic stroke. Stroke. 2010;41(5):910–915. doi:10.1161/STROKEAHA.109.577023

2. Feng W, Wang J, Chhatbar PY, et al. Corticospinal tract lesion load: An imaging biomarker for stroke motor outcomes. Ann Neurol. 2015;78(6):860–870. doi:10.1002/ANA.24510

3. Schulz R, Park CH, Boudrias MH, Gerloff C, Hummel FC, Ward NS. Assessing the Integrity of Corticospinal Pathways From Primary and Secondary Cortical Motor Areas After Stroke. Stroke. 2012;43(8):2248–2251. doi:10.1161/STROKEAHA.112.662619

4. Sperber C, Gallucci L, Mirman D, Arnold M, Umarova RM. Stroke lesion size – Still a useful biomarker for stroke severity and outcome in times of high-dimensional models. Neuroimage Clin. 2023;40:103511. doi:10.1016/J.NICL.2023.103511

5. Stinear CM, Barber PA, Smale PR, Coxon JP, Fleming MK, Byblow WD. Functional potential in chronic stroke patients depends on corticospinal tract integrity. Brain. 2007;130(Pt 1):170–180. doi:10.1093/BRAIN/AWL333

6. Schuch CP, Lam TK, Levin MF, et al. A comparison of lesion-overlap approaches to quantify corticospinal tract involvement in chronic stroke. J Neurosci Methods. 2022;376:109612. doi:10.1016/J.JNEUMETH.2022.109612

7. Boyd LA, Hayward KS, Ward NS, et al. Biomarkers of stroke recovery: Consensus-based core recommendations from the Stroke Recovery and Rehabilitation Roundtable. Int J Stroke. 2017;12(5):480–493. doi:10.1177/1747493017714176

8. Riley JD, Le V, Der-Yeghiaian L, et al. Anatomy of stroke injury predicts gains from therapy. Stroke. 2011;42(2):421–426. doi:10.1161/STROKEAHA.110.599340

9. Rondina JM, Park CH, Ward NS. Brain regions important for recovery after severe post-stroke upper limb paresis. J Neurol Neurosurg Psychiatry. 2017;88(9):737–743. doi:10.1136/JNNP-2016-315030

10. Rondina JM, Filippone M, Girolami M, Ward NS. Decoding post-stroke motor function from structural brain imaging. Neuroimage Clin. 2016;12:372–380. doi:10.1016/J.NICL.2016.07.014

11. Findlater SE, Hawe RL, Mazerolle EL, et al. Comparing CST Lesion Metrics as Biomarkers for Recovery of Motor and Proprioceptive Impairments After Stroke. Neurorehabil Neural Repair. 2019;33(10):848–861. doi:10.1177/1545968319868714

12. Lam TK, Binns MA, Honjo K, et al. Variability in stroke motor outcome is explained by structural and functional integrity of the motor system. Sci Rep. 2018;8(1):1–11. doi:10.1038/s41598-018-27541-8

13. Paul T, Wiemer VM, Hensel L, et al. Interhemispheric Structural Connectivity Underlies Motor Recovery after Stroke. Ann Neurol. 2023;94(4):785–797. doi:10.1002/ANA.26737

14. Esser F, Paul T, Rizor E, et al. Distinct Disconnection Patterns Explain Task-Specific Motor Impairment and Outcome After Stroke. Stroke. 2025;56(8):2210–2221. doi:10.1161/STROKEAHA.125.050929

15. Lin DJ, Cloutier AM, Erler KS, et al. Corticospinal Tract Injury Estimated from Acute Stroke Imaging Predicts Upper Extremity Motor Recovery after Stroke. Stroke. 2019;50(12):3569–3577. doi:10.1161/STROKEAHA.119.025898

16. Grafton ST, Fagg AH, Woods RP, Arbib MA. Functional anatomy of pointing and grasping in humans. Cereb Cortex. 1996;6(2):226–237. doi:10.1093/CERCOR/6.2.226

17. Carter AR, Patel KR, Astafiev S V., et al. Upstream dysfunction of somatomotor functional connectivity after corticospinal damage in stroke. Neurorehabil Neural Repair. 2012;26(1):7–19. doi:10.1177/1545968311411054

18. Kou N, Park CH, Seghier ML, Leff AP, Ward NS. Can fully automated detection of corticospinal tract damage be used in stroke patients? Neurology. 2013;80(24):2242–2245. doi:10.1212/WNL.0b013e318296e977

19. Paul T, Cieslak M, Hensel L, et al. The role of corticospinal and extrapyramidal pathways in motor impairment after stroke. Brain Commun. 2023;5(1):fcac301. doi:10.1093/BRAINCOMMS/FCAC301

20. Paul T, Cieslak M, Hensel L, et al. Corticospinal premotor fibers facilitate complex motor control after stroke. Ann Clin Transl Neurol. 2024;11(9):2439–2449. doi:10.1002/ACN3.52159

21. Beevor CE, Horsley VAH. III. An experimental investigation into the arrangement of the excitable fibres of the internal capsule of the bonnet monkey (macacus sinicus). Philos Trans R Soc Lond. 1890;(181):49–88. doi:10.1098/RSTB.1890.0003

22. Kunigk NG, Schone HR, Gontier C, et al. Motor somatotopy impacts imagery strategy success in human intracortical brain-computer interfaces. J Neural Eng. 2025;22(2):026004. doi:10.1088/1741-2552/ADB995

23. Penfield W, Boldrey E. Somatic motor and sensory representation in the cerebral cortex of man as studied by electrical stimulation. Brain. 1937;60(4):389–443. doi:10.1093/BRAIN/60.4.389

24. Olafson ER, Sperber C, Jamison KW, et al. Data-driven biomarkers better associate with stroke motor outcomes than theory-based biomarkers. Brain Commun. 2024;6(4):fcae254. doi:10.1093/BRAINCOMMS/FCAE254

25. Sperber C, Gallucci L, Arnold M, Umarova RM. The challenge of long-term stroke outcome prediction and how statistical correlates do not imply predictive value. Brain Commun. 2025;7(1):fcaf003. doi:10.1093/BRAINCOMMS/FCAF003

26. Wiemer VM, Paul T, Hensel L, et al. Structural reserve-dependent cortical rerouting of motor control after stroke. Brain. 2025:awaf434. doi:10.1093/BRAIN/AWAF434

27. Lyle RC. A performance test for assessment of upper limb function in physical rehabilitation treatment and research. Int J Rehabil Res. 1981;4(4):483–492. doi:10.1097/00004356-198112000-00001

28. Rorden C, Brett M. Stereotaxic Display of Brain Lesions. Behav Neurol. 2000;12(4):191–200. doi:10.1155/2000/421719

29. Grabner G, Janke AL, Budge MM, Smith D, Pruessner J, Collins DL. Symmetric Atlasing and Model Based Segmentation: An Application to the Hippocampus in Older Adults. Med Image Comput Comput Assist Interv. 2006;9(Pt 2):58–66. doi:10.1007/11866763_8

30. Chenot Q, Tzourio-Mazoyer N, Rheault F, et al. A population-based atlas of the human pyramidal tract in 410 healthy participants. Brain Struct Funct. 2019;224(2):599–612. doi:10.1007/S00429-018-1798-7

31. Archer DB, Vaillancourt DE, Coombes SA. A Template and Probabilistic Atlas of the Human Sensorimotor Tracts using Diffusion MRI. Cereb Cortex. 2018;28(5):1685–1699. doi:10.1093/CERCOR/BHX066

32. Lam TK, Cheung DK, Climans SA, et al. Determining Corticospinal Tract Injury from Stroke Using Computed Tomography. Can J Neurol Sci. 2020;47(6):775–784. doi:10.1017/CJN.2020.112

33. Benjamini Y, Hochberg Y. Controlling the False Discovery Rate: A Practical and Powerful Approach to Multiple Testing. J R Stat Soc: Series B Stat Methodol. 1995;57(1):289–300. doi:10.1111/J.2517-6161.1995.TB02031.X

34. Pedregosa F, Varoquaux G, Gramfort A, et al. Scikit-learn: Machine Learning in Python. J Mach Learn Res. 2011;12:2825–2830.

35. Garcea FE, Greene C, Grafton ST, Buxbaum LJ. Structural Disconnection of the Tool Use Network after Left Hemisphere Stroke Predicts Limb Apraxia Severity. Cereb Cortex Commun. 2020;1(1):1–20. doi:10.1093/TEXCOM/TGAA035

36. Sperber C, Karnath HO. Impact of correction factors in human brain lesion-behavior inference. Hum Brain Mapp. 2017;38(3):1692–1701. doi:10.1002/HBM.23490

37. Kasties V, Karnath HO, Sperber C. Strategies for feature extraction from structural brain imaging in lesion-deficit modelling. Hum Brain Mapp. 2021;42(16):5409–5422. doi:10.1002/HBM.25629

38. Cortes C, Vapnik V. Support-Vector Networks. Mach Learn. 1995;20:273-297. doi:10.1007/BF00994018

39. Michel V, Gramfort A, Varoquaux G, Eger E, Thirion B. Total variation regularization for fMRI-based prediction of behavior. IEEE Trans Med Imaging. 2011;30(7):1328–1340. doi:10.1109/TMI.2011.2113378

40. Winkler AM, Ridgway GR, Webster MA, Smith SM, Nichols TE. Permutation inference for the general linear model. Neuroimage. 2014;92:381–397. doi:10.1016/J.NEUROIMAGE.2014.01.060

41. Nichols TE, Holmes AP. Nonparametric permutation tests for functional neuroimaging: A primer with examples. Hum Brain Mapp. 2001;15(1):1–25. doi:10.1002/HBM.1058

42. Stinear CM, Lang CE, Zeiler S, Byblow WD. Advances and challenges in stroke rehabilitation. Lancet Neurol. 2020;19(4):348–360. doi:10.1016/S1474-4422(19)30415-6

43. Liew SL, Schweighofer N, Cole JH, et al. Association of Brain Age, Lesion Volume, and Functional Outcome in Patients With Stroke. Neurology. 2023;100(20):E2103–E2113. doi:10.1212/WNL.0000000000207219

44. Di Pino G, Pellegrino G, Assenza G, et al. Modulation of brain plasticity in stroke: a novel model for neurorehabilitation. Nat Rev Neurol. 2014;10(10):597–608. doi:10.1038/NRNEUROL.2014.162

45. Volz LJ, Rehme AK, Michely J, et al. Shaping Early Reorganization of Neural Networks Promotes Motor Function after Stroke. Cereb Cortex. 2016;26(6):2882–2894. doi:10.1093/CERCOR/BHW034

46. Puig J, Blasco G, Daunis-I-Estadella J, et al. Decreased corticospinal tract fractional anisotropy predicts long-term motor outcome after stroke. Stroke. 2013;44(7):2016–2018. doi:10.1161/STROKEAHA.111.000382

47. Byblow WD, Stinear CM, Barber PA, Petoe MA, Ackerley SJ. Proportional recovery after stroke depends on corticomotor integrity. Ann Neurol. 2015;78(6):848–859. doi:10.1002/ANA.24472

48. Newton JM, Ward NS, Parker GJM, et al. Non-invasive mapping of corticofugal fibres from multiple motor areas—relevance to stroke recovery. Brain. 2006;129(7):1844–1858. doi:10.1093/BRAIN/AWL106

49. Zarei M, Johansen-Berg H, Jenkinson M, Ciccarelli O, Thompson AJ, Matthews PM. Two-dimensional population map of cortical connections in the human internal capsule. J Magn Reson Imaging. 2007;25(1):48–54. doi:10.1002/JMRI.20810

50. Wilkins KB, Yao J, Owen M, Karbasforoushan H, Carmona C, Dewald JPA. Limited capacity for ipsilateral secondary motor areas to support hand function post-stroke. J Physiol. 2020;598(11):2153–2167. doi:10.1113/JP279377

