## Supplemental Material for "Predicting Motor Recovery After Stroke: Utility and Limits of Corticospinal Tract Biomarkers"

### Supplemental Methods

***Permutation-based contrast of voxel-wise feature weights between motor tasks***

To compare voxel-wise predictive relevance between basal and complex motor function, we computed contrasts between SVR-derived feature weights using a permutation-based approach.[40,41] For each permutation $k$, behavioral labels were randomly reassigned and SVR models were fitted separately for basal and complex motor function, yielding voxelwise weight maps $w_{v,k}^{\mathrm{perm,basal}}$and $w_{v,k}^{\mathrm{perm,complex}}$.These maps were subtracted to obtain a voxel-wise permutation difference map $\Delta w_{v,k}^{\mathrm{perm}}$. Across permutations, the mean $\mu(\Delta w_{v}^{\mathrm{perm}})$ and standard deviation $\sigma(\Delta w_{v}^{\mathrm{perm}})$ of the voxel-wise difference distribution were computed. For each voxel $v$, the observed difference in feature weights between models, $\Delta w_{v}^{\mathrm{true}}$, was expressed relative this permutation-derived difference distribution:

$$z_{v}^{\mathrm{diff}} = \frac{\Delta w_{v}^{\mathrm{true}} - \mu\left(\Delta w_{v}^{\mathrm{perm}}\right)}{\sigma\left(\Delta w_{v}^{\mathrm{perm}}\right)}.$$

This standardization provides a scale-invariant measure of relative differences between models and was used to facilitate comparison of voxel-wise contributions across models.

### Supplemental Figures

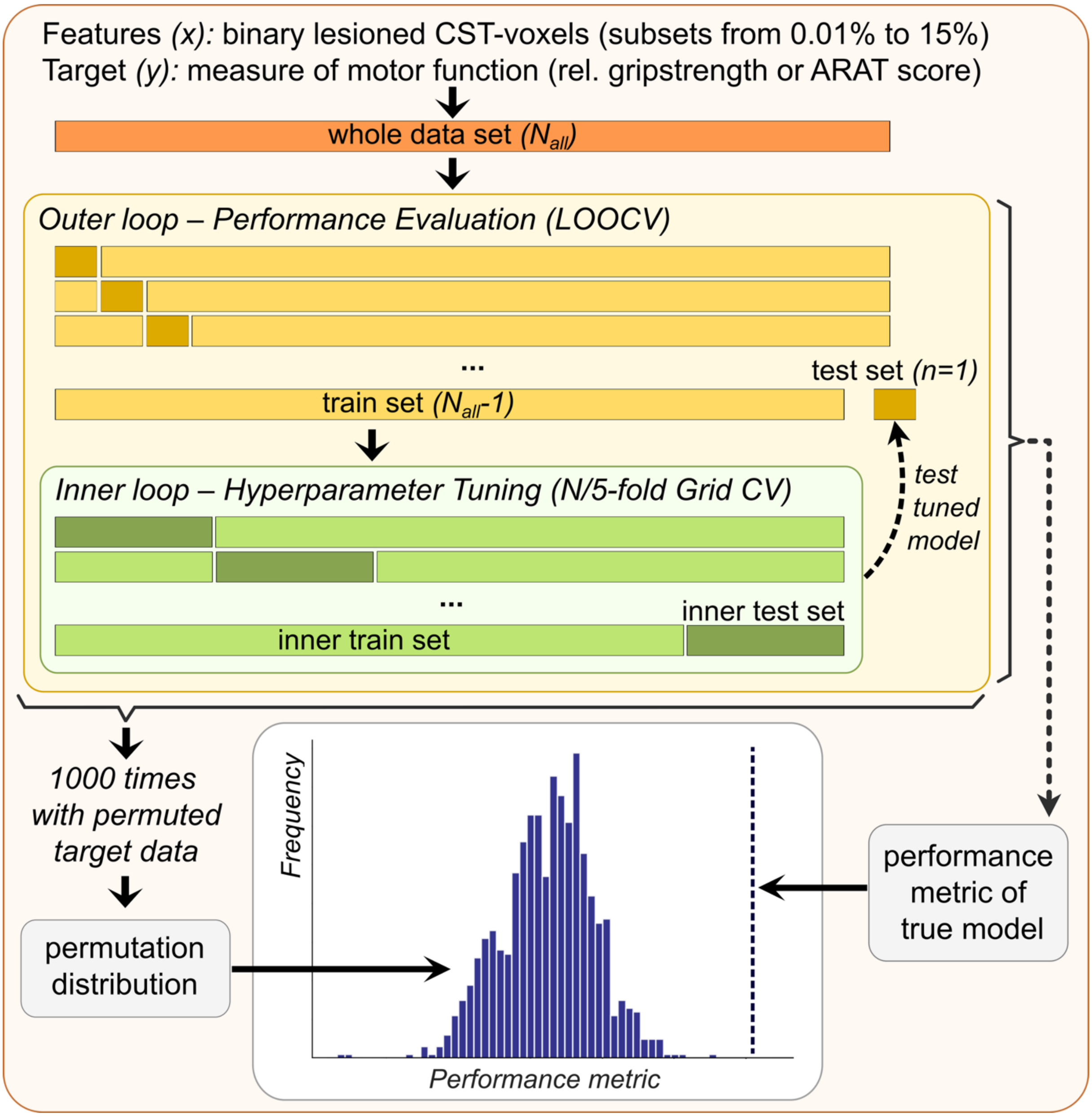

***Figure S1: Machine learning pipeline for voxel-wise prediction of upper extremity motor function from binary corticospinal tract (CST) lesion patterns.*** *For each analysis, input features (x) consist of binary lesioned CST voxels evaluated across manual feature selection thresholds ranging from 0.01% to 15%. The target variable (y) represents continuous scores of either basal (relative grip strength) or complex (Action Research Arm Test score) motor function. Model performance is evaluated in an outer leave-one-out cross-validation (LOOCV) loop, where a single patient is held out as an independent test set (dark yellow) for evaluation. The remaining train set (light yellow) is passed to an inner loop for hyperparameter optimization using a N/5-fold Grid Search cross-validation. Within the inner loop, parameters (e.g., SVR kernels) are tuned using the inner train set (light green), and performance is evaluated on the inner test set (dark green). The optimized model configuration is then deployed to predict the motor score of the completely unseen outer test case (dark yellow). To assess the statistical significance of the final cross-validated performance metrics, a permutation baseline distribution is generated by shuffling the true target data (y) relative to the input features and repeating the entire nested cross-validation pipeline 1,000 times. The performance metric of the true model (blue dashed vertical line) is then compared against this empirical permutation distribution (blue bars) to derive a non-parametric p-value.*

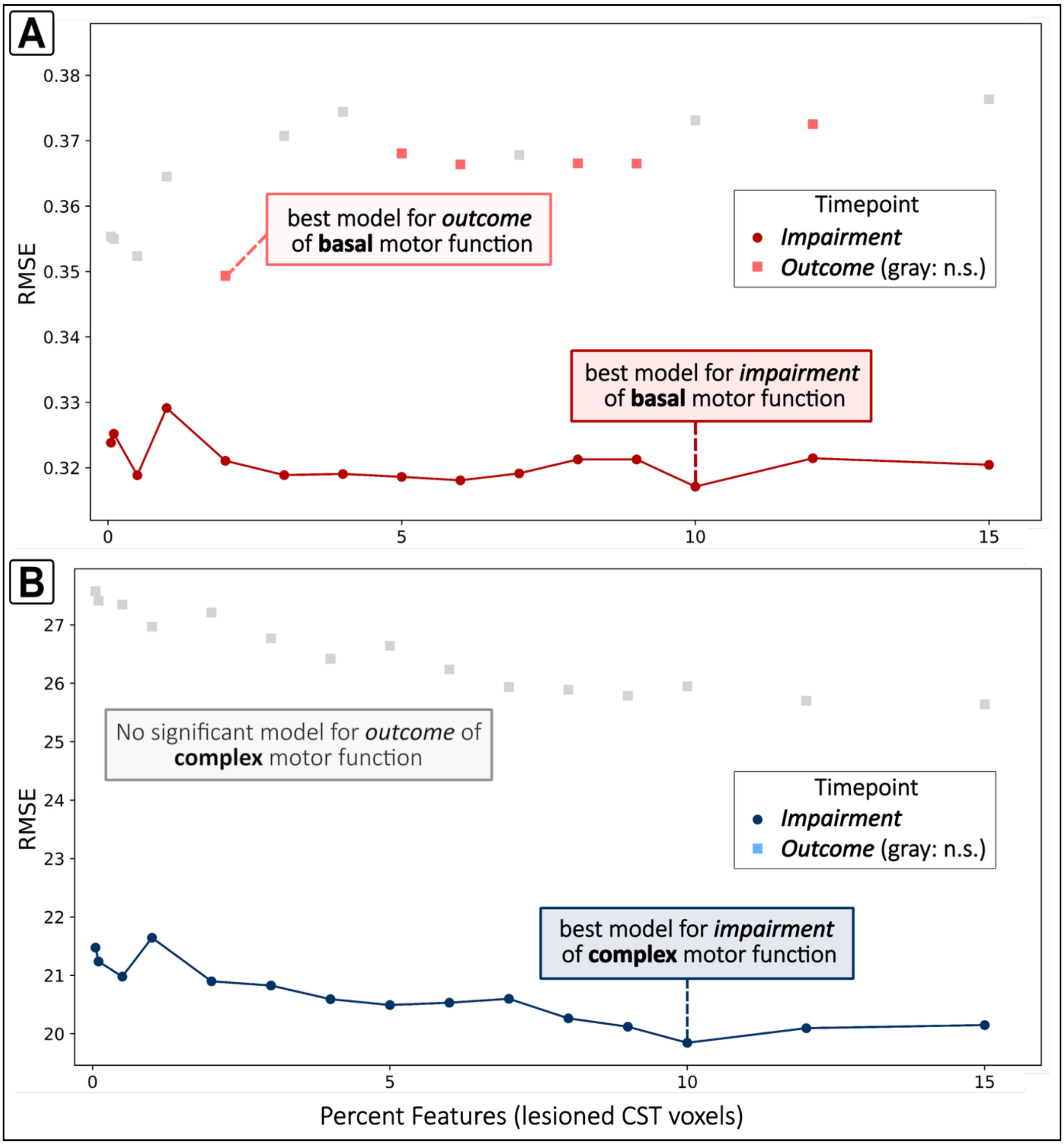


***Figure S2: Averaged Root mean squared error (RMSE) of SVR models across feature space.*** *Best performance for predicting impairment of basal (A) and complex (B) motor function was achieved when including 10 % of the most frequently lesioned CST voxels. For basal motor outcome, optimal performance was obtained with 2 % of voxels. No model significantly predicted complex motor outcome. Significant models were defined as those with* $R^2 > 0$ *and* $p < 0.05$.

### Supplemental Tables

| CST-lesion metric | Spearman correlation | | Cross-validated Spearman r | | Cross-validated $R^2$ | |
|---|---|---|---|---|---|---|
| **Complex impairment** | *r* | *p* | *r* | *p* | $R^2$ | *p* |
| Lesion volume | -0.23 | 0.015 | -0.20 | 0.172 | -0.01 | 0.128 |
| CST-lesion volume | -0.35 | <0.001 | 0.25 | 0.002 | 0.03 | 0.009 |
| raw CST-lesion overlap | -0.33 | <0.001 | 0.29 | 0.002 | 0.07 | 0.005 |
| weighted CST-lesion overlap | -0.38 | <0.001 | 0.34 | 0.002 | 0.12 | 0.004 |
| Feng-overlap | -0.37 | <0.001 | 0.32 | 0.002 | 0.06 | 0.004 |
| raw Zhu-overlap | -0.36 | <0.001 | 0.29 | 0.002 | 0.05 | 0.004 |
| weighted Zhu-overlap | -0.37 | <0.001 | 0.31 | 0.002 | 0.06 | 0.004 |
| **Complex outcome** | *r* | *p* | *r* | *p* | $R^2$ | *p* |
| Lesion volume | -0.13 | 0.222 | -0.17 | 0.107 | 0.03 | 0.025 |
| CST-lesion volume | -0.25 | 0.022 | 0.13 | 0.008 | 0.05 | 0.011 |
| raw CST-lesion overlap | -0.23 | 0.027 | 0.17 | 0.013 | 0.05 | 0.009 |
| weighted CST-lesion overlap | -0.27 | 0.015 | 0.22 | 0.010 | 0.08 | 0.004 |
| Feng-overlap | -0.25 | 0.022 | 0.20 | 0.007 | 0.09 | 0.005 |
| raw Zhu-overlap | -0.26 | 0.016 | 0.19 | 0.007 | 0.06 | 0.006 |
| weighted Zhu-overlap | -0.27 | 0.014 | 0.20 | 0.006 | 0.07 | 0.005 |

*continued from previous page*

| **CST-lesion metric** | **Spearman correlation** | | **Cross-validated Spearman *r*** | | **Cross-validated $R^2$** | |
|---|---|---|---|---|---|---|
| **Basal impairment** | ***r*** | ***p*** | ***r*** | ***p*** | $R^2$ | ***p*** |
| Lesion volume | -0.20 | 0.027 | -0.24 | 0.231 | -0.01 | 0.146 |
| CST-lesion volume | -0.34 | <0.001 | 0.25 | 0.004 | 0.03 | 0.009 |
| raw CST-lesion overlap | -0.34 | <0.001 | 0.30 | 0.002 | 0.07 | 0.005 |
| weighted CST-lesion overlap | -0.35 | <0.001 | 0.31 | 0.002 | 0.10 | 0.004 |
| Feng-overlap | -0.35 | <0.001 | 0.29 | 0.002 | 0.05 | 0.006 |
| raw Zhu-overlap | -0.36 | <0.001 | 0.30 | 0.002 | 0.05 | 0.006 |
| weighted Zhu-overlap | -0.37 | <0.001 | 0.31 | 0.002 | 0.06 | 0.005 |
| **Basal outcome** | ***r*** | ***p*** | ***r*** | ***p*** | $R^2$ | ***p*** |
| Lesion volume | -0.10 | 0.341 | -0.32 | 0.294 | -0.02 | 0.194 |
| CST-lesion volume | -0.25 | 0.020 | 0.10 | 0.013 | 0.00 | 0.065 |
| raw CST-lesion overlap | -0.23 | 0.027 | 0.15 | 0.015 | 0.00 | 0.047 |
| weighted CST-lesion overlap | -0.34 | 0.002 | 0.28 | 0.002 | 0.07 | 0.003 |
| Feng-overlap | -0.30 | 0.007 | 0.21 | 0.006 | 0.04 | 0.010 |
| raw Zhu-overlap | -0.28 | 0.013 | 0.16 | 0.011 | 0.01 | 0.034 |
| weighted Zhu-overlap | -0.30 | 0.008 | 0.20 | 0.009 | 0.02 | 0.019 |

***Table S1: Spearman correlation coefficients and cross-validated performance metrics of conventional CST-lesion metrics for predicting motor function.*** *For the cross-validated Spearman r and $R^2$, p-values derived from permutation testing are shown. All p-values were FDR-corrected for multiple comparisons.*

| SVR | Cross-validated Spearman *r* | | Cross-validated $R^2$ | |
|---|---|---|---|---|
| | *r* | *p* | $R^2$ | *p* |
| Complex impairment | 0.42 | <0.001 | 0.23 | <0.001 |
| Complex outcome | - | - | - | - |
| Basal impairment | 0.40 | <0.001 | 0.17 | <0.001 |
| Basal outcome | 0.33 | <0.001 | 0.13 | <0.001 |

***Table S2: Performance metrics of the best-performing SVR models predicting motor function.*** *Hyphens (-) indicate that no model achieved significant predictive performance for complex motor outcome. P-values were derived from permutation testing (1,000 permutations).*

| Complex | | Basal | |
|---|---|---|---|
| impairment<br>*10 % of features* | outcome | impairment<br>*10 % of features* | outcome<br>*2 % of features* |
| r = 0.77<br>p <0.001 | – | r = 0.90<br>p <0.001 | r = 0.85<br>p < 0.001 |

***Table S3: Spearman correlation coefficients for feature weights of best nonlinear SVR model and linear SVR model.*** *Feature weights were highly correlated for selected models indicating that both, linear and nonlinear models rely on similar support vectors and that the superior model performance of nonlinear models was likely not due to overfitting. Spearman correlation was assessed due to non-normality of distribution in the Shapiro-Wilk-Test.*

STROBE Statement—Checklist of items that should be included in reports of ***cohort studies***

| | Item No | Recommendation | Page No |
|---|---|---|---|
| **Title and abstract** | 1 | (*a*) Indicate the study's design with a commonly used term in the title or the abstract | p.1, ll.8-13 |
| | | (*b*) Provide in the abstract an informative and balanced summary of what was done and what was found | p.1, ll.8-22 |
| **Introduction** | | | |
| Background/rationale | 2 | Explain the scientific background and rationale for the investigation being reported | p.2,l.35- p. 3 l.80 |
| Objectives | 3 | State specific objectives, including any prespecified hypotheses | p.3,l.80-p.4,l.98 |
| **Methods** | | | |
| Study design | 4 | Present key elements of study design early in the paper | p.5,l.104-p.6,l.128 |
| Setting | 5 | Describe the setting, locations, and relevant dates, including periods of recruitment, exposure, follow-up, and data collection | p.5,l.104-p.6,l.128 |
| Participants | 6 | (*a*) Give the eligibility criteria, and the sources and methods of selection of participants. Describe methods of follow-up | p.5,l.104-p.6,l.128 |
| | | (*b*) For matched studies, give matching criteria and number of exposed and unexposed | N/A |
| Variables | 7 | Clearly define all outcomes, exposures, predictors, potential confounders, and effect modifiers. Give diagnostic criteria, if applicable | p.5,l.115-p.12,l.251 |
| Data sources/ measurement | 8* | For each variable of interest, give sources of data and details of methods of assessment (measurement). Describe comparability of assessment methods if there is more than one group | p.5,l.115-p.12,l.249 |
| Bias | 9 | Describe any efforts to address potential sources of bias | p.11,l.217-p.12,l.249 p.24,l.360-371 |
| Study size | 10 | Explain how the study size was arrived at | p.6,ll.125-128 |
| Quantitative variables | 11 | Explain how quantitative variables were handled in the analyses. If applicable, describe which groupings were chosen and why | p.7,l.146-p.12,l.249 |
| Statistical methods | 12 | (*a*) Describe all statistical methods, including those used to control for confounding | p.7,l.146-p.12,l.249 |
| | | (*b*) Describe any methods used to examine subgroups and interactions | N/A |
| | | (*c*) Explain how missing data were addressed | N/A |
| | | (*d*) If applicable, explain how loss to follow-up was addressed | N/A |
| | | (*e*) Describe any sensitivity analyses | N/A |
| **Results** | | | |
| Participants | 13* | (a) Report numbers of individuals at each stage of study—eg numbers potentially eligible, examined for eligibility, confirmed eligible, included in the study, completing follow-up, and analysed | p.5,ll.104-106, p.13, ll.251-254 |
| | | (b) Give reasons for non-participation at each stage | p.6,ll.125-128 |
| | | (c) Consider use of a flow diagram | N/A |
| Descriptive data | 14* | (a) Give characteristics of study participants (eg demographic, clinical, social) and information on exposures and potential confounders | p.13,ll.251-260, p.24,ll.360-371 |

| | | | |
|---|---|---|---|
| | | (b) Indicate number of participants with missing data for each variable of interest<br><br>(c) Summarise follow-up time (eg, average and total amount) | p.6,ll.125-128<br>p.13, ll. 251-254<br>p.13, ll. 251-254 |
| Outcome data | 15* | Report numbers of outcome events or summary measures over time | pp.13,14, ll.251-260, Fig 2 |

| | | | |
|---|---|---|---|
| Main results | 16 | (*a*) Give unadjusted estimates and, if applicable, confounder-adjusted estimates and their precision (eg, 95% confidence interval). Make clear which confounders were adjusted for and why they were included<br>(*b*) Report category boundaries when continuous variables were categorized<br>(*c*) If relevant, consider translating estimates of relative risk into absolute risk for a meaningful time period | p.13,l.251-p.19, l.338; Fig 3-6<br>N/A<br>N/A |
| Other analyses | 17 | Report other analyses done—eg analyses of subgroups and interactions, and sensitivity analyses | N/A |
| **Discussion** | | | |
| Key results | 18 | Summarise key results with reference to study objectives | p.30 l.513-p.31,l.531 |
| Limitations | 19 | Discuss limitations of the study, taking into account sources of potential bias or imprecision. Discuss both direction and magnitude of any potential bias | p.29.,l.494-p.30,l.511 |
| Interpretation | 20 | Give a cautious overall interpretation of results considering objectives, limitations, multiplicity of analyses, results from similar studies, and other relevant evidence | p.25,l.372-p.30,l.492, p.30 l.513-p.31,l.531 |
| Generalisability | 21 | Discuss the generalisability (external validity) of the study results | p.31, ll.527-531 |
| **Other information** | | | |
| Funding | 22 | Give the source of funding and the role of the funders for the present study and, if applicable, for the original study on which the present article is based | p.32, ll.536-539 |

*Give information separately for exposed and unexposed groups.

**Note:** An Explanation and Elaboration article discusses each checklist item and gives methodological background and published examples of transparent reporting. The STROBE checklist is best used in conjunction with this article (freely available on the Web sites of PLoS Medicine at http://www.plosmedicine.org/, Annals of Internal Medicine at http://www.annals.org/, and Epidemiology at http://www.epidem.com/). Information on the STROBE Initiative is available at http://www.strobe-statement.org.
